# Effects of Pycnogenol^®^ in post-COVID-19 condition (PYCNOVID): A single-center, placebo controlled, quadruple-blind, randomized trial

**DOI:** 10.1101/2025.06.12.25329489

**Authors:** Julia Kopp, Lisa Künzi, Sonja Rüegg, Julia Braun, Manuela Rasi, Alexia Anagnostopoulos, Milo A. Puhan, Jan Sven Fehr, Thomas Radtke

**Affiliations:** Epidemiology, Biostatistics and Prevention Institute, University of Zurich, Hirschengraben 84, 8001 Zürich, Switzerland

## Abstract

**Background:** Post-COVID-19 condition (PCC) remains a public health challenge with limited treatment options.

**Objective:** To evaluate the effects of Pycnogenol^®^, a French maritime pine bark extract with anti-inflammatory and antioxidative properties, versus placebo on patient-reported health status in individuals with PCC.

**Design:** Single-center, placebo-controlled, quadruple-blind, randomized trial. ClinicalTrials.gov, NCT05890534

**Setting:** German-speaking Switzerland.

**Participants:** Adults with PCC with at least one of the following symptoms: fatigue, post-exertional malaise, dyspnea, or brain fog.

**Intervention:** Daily dose of 200 mg Pycnogenol^®^ (50 mg capsules four times daily) or placebo for 12 weeks.

**Measurements:** The primary outcome was self-reported health status (EQ-VAS). Secondary outcomes included symptoms, quality of life, exercise capacity, physical activity and blood biomarkers.

**Results:** Between June 14, 2023, and July 5, 2024, 153 participants were randomized to receive Pycnogenol^®^ (n=75) or placebo (n=78). Participants reported persisting symptoms for a median duration of 101 weeks. Unadjusted median EQ-VAS increased by 5.4 units and 7.9 units in the in the Pycnogenol^®^ and placebo groups, respectively, with no difference observed between the groups (β=0.54, 95% CI -3.45 to 4.54, p=0.79). 31 adverse events occurred in the Pycnogenol^®^ group (11 probably related) and 18 in the placebo group (9 probably related). Three serious adverse events were reported, all unrelated to the study products. No between group differences were observed in secondary endpoints.

**Limitation:** The results may not be generalizable to individuals newly experiencing PCC.

**Conclusion:** Pycnogenol^®^ (200 mg daily) did not improve health status compared to placebo over 12 weeks in PCC. Both groups showed clinically relevant improvements, suggesting non-therapeutic effects.

**Primary funding source:** Horphag Research, Switzerland.

## Introduction

Many individuals continue to experience long-term effects of Severe Acute Respiratory Syndrome Coronavirus 2 (SARS-CoV-2) infection. While most recoveries are uncomplicated, some individuals experience persistent and significant sequelae long after the acute phase of illness (1,2). To characterize the broad spectrum of physical and mental symptoms that may persist or emerge three months or more following SARS-CoV-2 infection, the World Health Organization (WHO) introduced the term “post-COVID-19 condition” (PCC) (3,4). The symptoms can affect multiple organ systems and are often marked by fluctuating periods of improvement followed by relapses (1). Population-based studies report a prevalence of about 20% in individuals with confirmed infection and 5-10% among all infected adults (5,6). Findings from our population-based cohort study (Zurich SARS-CoV-2 Cohort) offer valuable insights into symptom patterns and the natural progression of PCC (7,8). In this cohort study, about 17% of persons with confirmed infection reported having not yet recovered at 24 months post-infection with about 10% reporting mild, 4% moderate, and 2% severe impairment (7). Fatigue and post-exertional malaise (PEM) are the most common symptoms along with dyspnea and concentration difficulties (brain fog) (9,10). The pathogenesis of PCC is thought to be multifactorial and complex, with several hypotheses proposed to explain the persistence of symptoms and health complication including viral persistence, immune dysregulation, hypercoagulability and platelet activation, microvascular dysfunction, inflammation, mitochondrial dysfunction, and gut dysbiosis (11).

As of the end of Mai 2025, approximately 400 interventional PCC studies were registered on ClinicalTrials.gov. These studies mostly focus on physical training and rehabilitation, while only few explore pharmaceutical drugs, some with significant risks such as immune suppression, metabolic side effects, gastrointestinal symptoms and other unknown long-term effects (12–14). Recently published, placebo-controlled randomized-controlled trials (RCTs) on antiviral therapy (nirmatrelvir-ritonavir) or nutritional supplements (lithium aspartate) showed no improvements in persisting symptoms compared to placebo. To date, most RCTs investigating pharmaceutical treatments have not found benefits over placebo in improving fatigue, cognitive dysfunction, shortness of breath, and other symptoms associated with PCC (15–19). In a double-blind RCT, Lau et al. found that SIM01, a synbiotic formulation, was well tolerated and improved multiple PCC symptoms and gut microbiota composition after six months compared to placebo (20). While a single treatment is unlikely to solve the complexity of symptoms in PCC, these data highlight the importance to study treatment options with a favorable benefit-risk ratio (12–14).

Pycnogenol^®^ is a standardized extract from French maritime pine bark, primarily composed of procyanidins (flavonoid polymers) and their main monomers catechin and epicatechin, as well as phenolic acids (21). It is known to have few side effects and has a long history on the market (22,23). The product has antioxidative, anti-inflammatory, and antiproliferative properties and has been shown to improve vascular endothelial function, probably due to an upregulation of endothelial nitric oxide synthase activity (23–25). Daily intake of 150mg of Pycnogenol^®^ over 12 weeks improved endothelial function, inflammatory and antioxidative biomarkers compared to placebo in individuals recovering from COVID-19 (22). Consequently, it might be an interesting candidate to improve health status and symptoms in people with PCC.

We therefore conducted an RCT to evaluate the efficacy of Pycnogenol^®^ versus placebo on the health status of individuals with PCC.

## Methods

### Study design

The study was a single-center, placebo-controlled, quadruple-blind, parallel design randomized superiority trial of 12 weeks duration, in which participants were randomly allocated to receiving either Pycnogenol^®^ or placebo. The trial was conducted at the University of Zurich (UZH), Switzerland between June 2023 and November 2024. The trial was approved by the ethics committee of the Canton of Zurich, Switzerland (Kantonale Ethik Kommission Zürich; BASEC-Nr. 2022-01967) and the study protocol has been published previously (26). We actively involved six people with lived experience in the study planning process, and the selection of the primary endpoint to capture their daily symptom burden. Moreover, they provided valuable feedback with respect to the organization of the study visits (27).

This study is reported according to the CONSORT guidelines for reporting randomized controlled parallel-design studies (28).

### Participants

We included participants aged 18 years or older who were fluent in German, without anticipated changes in medication, had a confirmed SARS-CoV-2 infection (verified through Polymerase Chain Reaction (PCR), rapid antigen test, or a physician diagnosis of PCC), were not suffering from untreated comorbidities, and had at least one of the following persisting symptoms: fatigue, cognitive impairment (“brain fog”), dyspnea, or post-exertional malaise (PEM). Participants were recruited through various channels including the Altea network, the Long COVID Citizen Science Board, the Facebook group of the patient organization Long COVID Schweiz, and advertisements in public transport systems in the city of Zurich and its surrounding metropolitan area. Furthermore, online flyers were shared with different stakeholders, primarily hospitals, to increase awareness of the study. Written informed consent was obtained from all participants.

### Randomization and masking

Participants were randomly allocated to either receive a daily dose of 200mg Pycnogenol^®^ or placebo for 12 weeks using stratified block randomization with a 1:1 allocation ratio. Stratification was done for the presence of chronic symptomatic disease(s) and duration of PCC (≤ six months versus > six months). We used six different randomization groups (A, B, C, D, E, F) with three representing placebo and three Pycnogenol^®^. The entire study team remained blinded to group assignment until the study was completed. The list of random numbers (block size of six representing the six randomization groups) was generated by a senior biostatistician not involved in the trial using the package blockrand with the software R (29), and implemented into the REDCap (Research Electronic Data Capture, Vanderbilt University, USA) database by the same person. Computer-based randomization was performed during the baseline visit within the REDCap database allowing for complete concealment of the allocation sequence for study personnel (30).

Trial participants, team of investigators, outcome assessors, and data analysts including laboratory personnel were blinded. study products were identical in appearance and appropriately labeled, and the de-identification code was only accessible to the principal investigator. A team of researchers from our institute, not involved in the trial, was responsible for study product labeling. Study products were manufactured in a Good Manufacturing Practice-accredited facility and stored at our study center under controlled conditions (Supplementary Table 9).

### Assessments and procedures

The study comprised four visits with a 12-week intervention phase (Supplementary Figure 1). Prior to the screening visit, participants received an online questionnaire asking for personal contact details, demographics, socioeconomic status, medical history, symptoms, and self-rated health status. The screening visit included obtaining informed consent and various assessments to clarify eligibility (Supplementary Figure 1). Approximately two weeks later, participants were asked to complete an online questionnaire, including self-assessments of persisting symptoms, validated instruments assessing fatigue, anxiety and depression, and health-related quality of life including health status. Additionally, physical activity was measured before the first study visit. The baseline visit included various assessments (Supplementary Figure 1) and the randomization process. Following randomization, participants received their assigned study products, either 200mg Pycnogenol^®^ (50mg per capsule, taken as 2 capsules in the morning an evening each) or placebo with identical appearance. At the end of the baseline visit, participants were instructed to fill-out a weekly symptom diary tracking PCC symptoms and their severity. To ensure the validity of our data and promote adherence to the study protocol, participants attended a brief study visit (follow-up 1) six weeks after randomization (exchange of study products), plus two follow-up phone calls at two- and nine-weeks post-randomization. After the 12-week intervention, the final study visit (follow-up 2) was conducted, which included the same online questionnaires and assessments as at baseline (Supplementary Figure 1). After completion of this visit, participants were invited to complete an online survey about their experience during the study. All questionnaires were completed online using REDCap. To ensure compliance and systematic monitoring of all adverse events (AEs), the study team stayed in contact with all participants through E-mail, phone calls and follow-up visits. AEs were classified into five categories: gastrointestinal, dermatological, physical-activity-related, respiratory, and other complaints. The trained study team assessed participant safety and causality. Serious adverse events (SAEs) related to the intervention could lead to early trial termination. In addition, we assessed whether the study visits posed a burden on the participants. At each study visit, starting with the baseline visit, we asked participants whether they experienced a worsening of their symptoms within 3 days after the previous study visit.

### Outcomes

The primary outcome was the self-reported health status assessed through the online questionnaires using the EuroQol visual analogue scale (EQ-VAS) over seven consecutive days before baseline and follow-up 2.(31) The EQ-VAS is numbered from 0 to 100 with 0 representing the “worst imaginable health” and 100 the “best imaginable health” (32). EQ-VAS values were analyzed for participants who completed at least four of seven EQ-VAS assessments before baseline and follow-up 2.

Some secondary outcomes were assessed prior to the baseline and follow-up 2 visits except functional exercise capacity, cognitive function assessment, and blood biomarkers. Fatigue was assessed using the 13-item Functional Assessment of Chronic Illness Therapy – Fatigue (FACIT-Fatigue) instrument (33–35), and the fatigue domain of the Chronic Respiratory Questionnaire (CRQ). Dyspnea, emotional function, and mastery were assessed with the CRQ (36–38). Depression and anxiety were assessed using the Hospital Anxiety and Depression Scale (HADS) (39). Quality of life was assessed using the EuroQol EQ-5D-5L with the Dutch value set to calculate EQ-5D-5L index scores. The questionnaire includes five dimensions with five levels of severity (32,40). Cognitive function was assessed using the Montreal Cognitive Assessment Test (MoCa) with a cut-off score <26 for impairment. The standard correction for years of education was applied (41–45). In addition to validated questionnaires, self-reported symptoms, including a severity grading on a 5-point Likert scale ("not bad at all", “mild”, “moderate”, “severe”, and "very severe"), were assessed before the visits and weekly throughout the 12-week intervention phase using a paper symptom diary. Self-reported dyspnea was assessed using three items: shortness of breath, breathing difficulties at rest, and breathing difficulties during exertion. Self-reported brain fog was evaluated based on two items: concentration difficulties and memory problems. Functional exercise capacity was measured using the 30s sit-to-stand (30s STS) test (46–49) and habitual physical activity was measured with a triaxial accelerometer (ActiGraph wGT3x-BT, Pensacola, FL, USA) for eight consecutive days before baseline and follow-up 2 (50,51). Blood biomarkers for endothelial health, inflammation, coagulation and platelet function, oxidative stress, and liver and kidney function included blood count, D-dimers, international normalized ratio (INR), activated partial thromboplastin time (aPTT), soluble CD40 ligand (sCD40L), soluble platelet selectin (sP-selectin), soluble thrombomodulin (s-TM), von Willebrand factor (vWF) antigen, syndecan-1, soluble vascular cell adhesion molecule-1 (sVCAM-1), C-reactive protein (CRP), interleukin-6 (IL-6), total antioxidative capacity (TAC), creatinine, and the liver enzymes aspartate aminotransferase (ASAT), alanine aminotransferase (ALAT) and gamma glutamyltransferase (γ-GT). Further information on methods and instruments including references can be found in the online supplementary file.

### Sample size

Sample size calculations were based on data from the Zurich SARS-CoV-2 Cohort, a population-based prospective cohort study reflecting the natural course of PCC without treatment.(52) Among 1543 participants in this cohort, 208 (13.5%) reported at least one persisting symptom – such as fatigue, PEM, dyspnea, or brain fog – six months after PCR-confirmed SARS-CoV-2 infection. The mean (SD) health status, assessed using the EQ-VAS (31) on a 0-100 scale (0 = worst health status; 100 = best health status), was 73.4 ± 16.5. Based on this data and the minimal important difference (MID) of 8 units for the EQ-VAS (52), a sample size of 136 participants (68 per group) was needed to provide 80% power to detect a difference of 8 units between the Pycnogenol^®^ and placebo groups at 12 weeks. To account for an anticipated dropout rate of 10%, the study aimed to enroll a total of 150 participants.

### Statistical analysis

The primary analysis was based on the intention-to-treat (ITT) principle. Per protocol analyses were done as sensitivity analysis. All randomized trial participants having received at least one dose of the study product were considered for the analysis. Baseline characteristics of the study participants were summarized with numbers and percentages for qualitative variables, and median and 25^th^-75^th^ percentiles for quantitative variables. The blood biomarkers have a skewed distribution. We log-transformed these data to achieve a distribution closer to normal. A per-protocol analysis was executed to compare adherent versus non-adherent participants. Adherence was defined as taking more than 70% of the prescribed capsules throughout the study (26).

Both, primary and secondary endpoints were analyzed with multivariable linear regression models adjusted for baseline values and presence of chronic symptomatic disease at baseline (stratification factor). Missing values for two participants in the EQ-VAS follow-up 2 were imputed using multiple imputation by chained equations (MICE) with predictive mean matching, based on 20 imputed datasets (mice package, R version 4.2.2). The models for physical activity were additionally adjusted for accelerometer wear time. Changes in symptoms at follow-up 2 were restricted to those relevant for inclusion into the trial such as dyspnea, fatigue, concentration difficulties (brain fog) and PEM and were analyzed using logistic regression models adjusted for baseline values and chronic symptomatic disease. Changes in symptom severity over time were analyzed using a generalized linear mixed-effects model (GLMM) with a binomial family (exploratory analysis). All analyses were conducted in R version 4.2.2. A data monitoring committee was responsible for ensuring data quality during and after the study. The external monitoring was conducted by MEDICRO GmbH, Petersaurach, Germany. This trial was registered on ClinicalTrials.gov, NCT05890534.

## Results

Between June 14, 2023, and July 5, 2024, 418 individuals were pre-screened for eligibility. Among them, 248 did not fulfil the eligibility criteria leaving 170 individuals who were invited for screening. Of those, 153 met the eligibility criteria, consented to participate, and were subsequently randomized (Figure 1). A total of 78 participants were assigned to the placebo group, and 75 were assigned to the Pycnogenol^®^ group. Table 1 summarizes the baseline characteristics of the trial participants, stratified by randomization group. The median time since participants’ SARS-CoV-2 infection causing persisting symptoms was on average approximately 100 weeks. At baseline, the number and percentage of participants with fatigue (FACIT-Fatigue, score <34), depression or anxiety (HADS, score ≥7), and cognitive impairment (MoCA, score <26) were as follows: fatigue – 126 (82.9%); anxiety – 61 (39.9%); depression - 69 (45.1%); and cognitive impairment – 19 (12.8%). In addition, perceived health status (EQ-VAS) was remarkably low. Supplementary Table 1 summarizes the administered medications and therapies reported by study participants at baseline.

**Figure 1.**
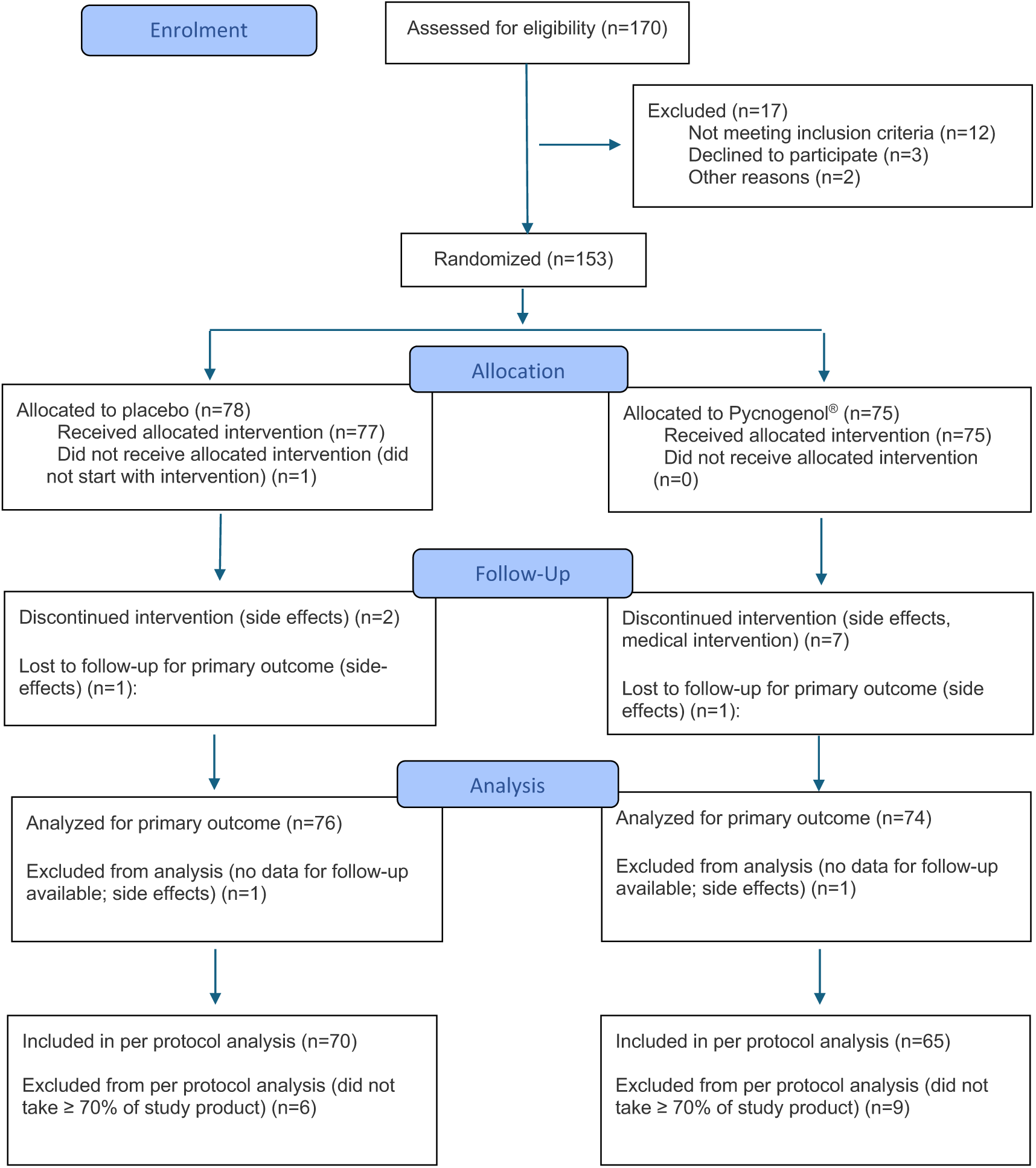
PYCNOVID flow diagram showing the progress of the participants through the randomized trial.

**Table 1.** Participant characteristics at baseline.

| Characteristics | Placebo (n=78) | Pycnogenol® (n=75) |
| --- | --- | --- |
| **Female** | 56 (71.8) | 59 (78.7) |
| **Age, years** | 43.5 [33.0 - 55.0] | 45.0 [36.0 - 54.0] |
| **Smoking status** |  |  |
| Never | 46 (59.0) | 52 (69.3) |
| Former | 26 (33.3) | 17 (22.7) |
| Current | 6 (7.7) | 6 (8.0) |
| **Hospitalized for SARS-CoV-2 infection** | 3 (3.8) | 3 (4.0) |
| **Vaccinated against SARS-CoV-2** | 73 (93.6) | 70 (93.3) |
| **Time since onset of symptoms, weeks** | 97.3 [73.6 – 131.7] | 106.6 [74.9 – 147.1] |
| **Spirometry** |  |  |
| FVC < LLN | 4 (5.1) | 4 (5.3) |
| FEV<sub>1</sub> < LLN | 8 (10.3) | 10 (13.3) |
| FEV<sub>1</sub>/FVC < LLN | 7 (9.0) | 5 (6.7) |
| **Chronic symptomatic disease<sup>#</sup>** | 24 (30.8) | 24 (32.0) |
| **Comorbidities** |  |  |
| Cardiovascular Disease | 2 (2.6) | 5 (6.7) |
| Hypertension | 4 (5.1) | 8 (10.7) |
| Diabetes | 1 (1.3) | 2 (2.7) |
| Obesity | 15 (19.2) | 15 (20.0) |
| Chronic Respiratory Disease | 15 (19.2) | 7 (9.3) |
| Chronic Kidney Disease | 3 (3.8) | 0 (0) |
| Autoimmune Disease | 3 (3.8) | 5 (6.7) |
| Psychiatric Disease | 8 (10.3) | 12 (16.0) |
| **FACIT-Fatigue** | 22.0 [15.0 – 30.0] | 24.0 [19.2 – 29.8] |
| **CRQ domains** |  |  |
| Fatigue | 3.8 [3.0 – 4.5] | 3.2 [2.8 – 4.0] |
| Dyspnea | 6.0 [4.8 – 6.8] | 6.0 [5.2 – 6.6] |
| Emotional | 4.6 [4.0 – 5.3] | 4.6 [4.0 – 5.2] |
| Mastery | 6.0 [5.0 – 6.2] | 6.0 [5.2 – 6.2] |
| **HADS domains** |  |  |
| Anxiety | 5.0 [3.0 – 8.0] | 5.0 [3.0 – 8.0] |
| Depression | 6.0 [4.0 – 9.0] | 6.0 [4.0 – 8.0] |
| **MoCA Score** | 28.0 [26.8 – 29.0] | 28.0 [27.0 – 29.0] |
| **EQ-VAS daily\*** | 48.2 [34.8 – 65.2] | 51.4 [40.1 – 63.1] |

Data are n (%) or median [IQR] unless otherwise specified; Obesity defined as body mass index > 30kg/m<sup>2</sup>; FEV<sub>1</sub>: Forced expiratory volume in one second; FVC: Forced vital capacity; LLN: Lower limit of normal (i.e., corresponds to a z-score value < -1.645); FACIT-Fatigue: Functional Assessment of Chronic Illness Therapy – Fatigue; CRQ: Chronic Respiratory Questionnaire; HADS: Hospital Anxiety and Depression Score; MoCA: Montreal Cognitive Assessment; EQ-VAS: EuroQol Visual Analogue Scale. \*Median values were calculated from the mean value from daily EQ-VAS data for each individual participant at baseline. # Stratification variable including symptomatic chronic respiratory disease, psychiatric disorders, cardiovascular disease, autoimmune disease, obesity, and diabetes. Note: Numbers differ from those reported for comorbidities as only symptomatic diseases were considered for stratified randomization.

During the study, two participants from the placebo group and three from the Pycnogenol^®^ group stopped their study participation, leaving 148 participants who completed all study visits. Two participants of the Pycnogenol^®^ group who stopped with the study products provided data for the primary outcome (EQ-VAS) and were included in the intention-to-treat (ITT) analysis. Furthermore, we imputed EQ-VAS values for two participants who did not fill-out the primary outcome at follow-up 2, resulting in a total of 152 individuals with primary outcome data. One participant from the placebo group did not start with study product and did therefore not qualify for the ITT analysis (Figure 1). Ultimately, complete data for all analyses were available for 146 participants (95% of the total sample) due to missing data in one baseline questionnaire and one follow-up 2 questionnaire.

Figure 2 shows the changes in mean EQ-VAS scores (primary endpoint) from baseline to follow-up 2 for the two groups separately. Each boxplot represents the distribution of individual scores, with jittered dots showing individual mean data points (i.e., mean of all available EQ-VAS data from baseline and follow-up 2). Overall, a total of 1015 EQ-VAS datapoints (on average 6.7 per participant) at baseline and 1003 EQ-VAS datapoints (on average 6.7 per participant) at follow-up 2 were available for the analysis. In the Pycnogenol^®^ group, the unadjusted median EQ-VAS increased by 5.4 units and 7.9 units in the placebo group. At 12 weeks, there was no significant difference in EQ-VAS values between the two groups (β=0.54, 95% CI -3.45 to 4.54, p=0.79). We conducted multiple imputation for the missing values; however, this did not alter the outcome. Mean baseline EQ-VAS values were strongly associated with the mean follow-up 2 EQ-VAS values (β=0.91, 95% CI 0.80 to 1.03, p<0.001), indicating that higher baseline values were predictive of higher follow-up 2 values. Supplementary Figure 2 presents boxplots of EQ-VAS values at baseline and at follow-up 2 across seven time points, stratified by randomization groups. Both groups showed similar distributions at baseline, with a trend towards higher EQ-VAS values at follow-up 2. The plot also highlights the large variability across time points in both groups.

**Figure 2.**
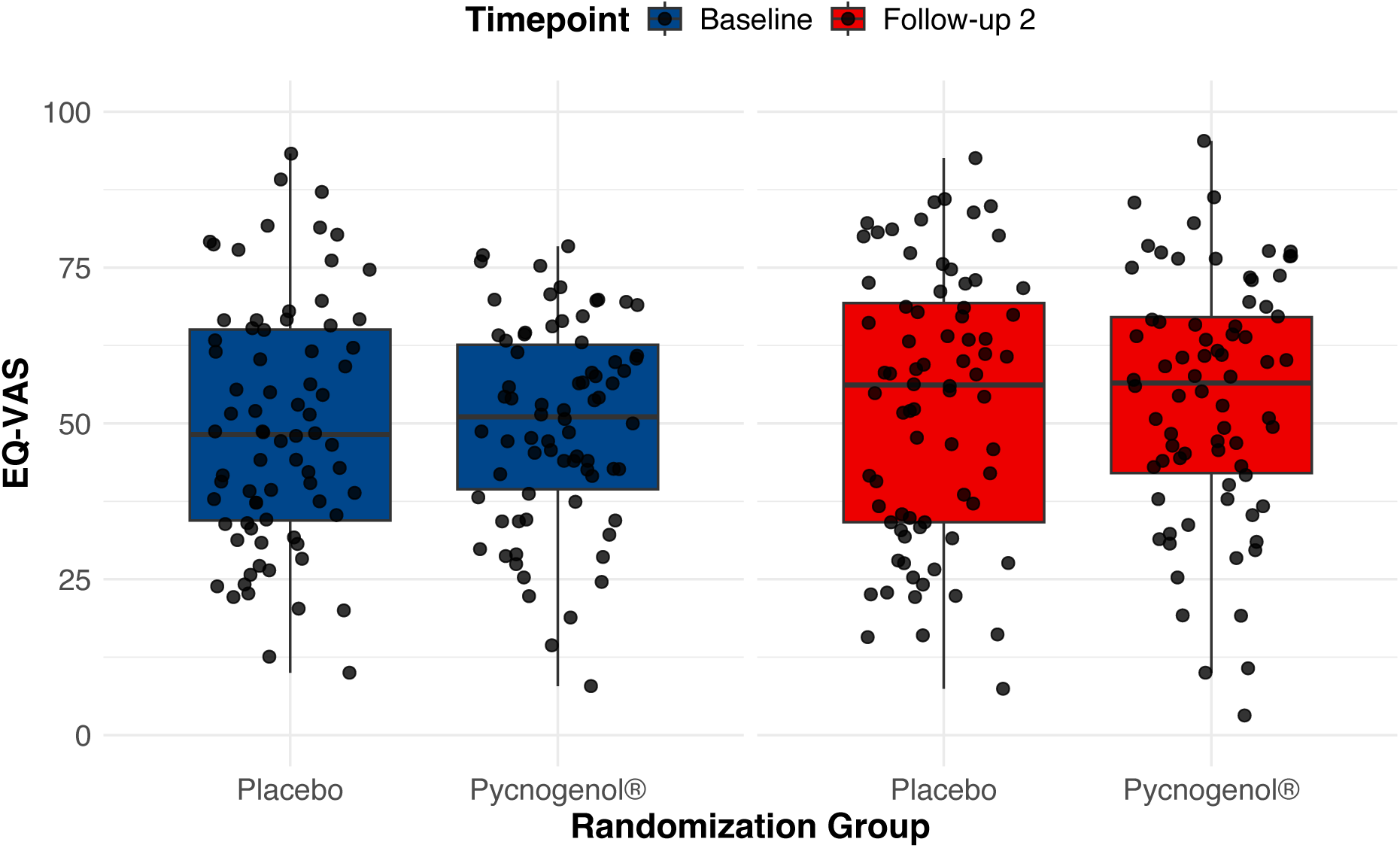
Box and Whiskers plot showing EQ-VAS scores at baseline and follow-up by randomization group (N=150). Median values are annotated in each box. Raw, unadjusted data are displayed as individual datapoints, with each participant’s value representing the mean of up to seven daily measurements per timepoint.

### Secondary Outcomes

Table 2 summarizes the results of the secondary outcomes. The time spent sedentary (β =-22.47 min.day-1, 95% C -40.27 to -4.67, p=0.01) and self-reported dyspnea (β=-1.08, 95% CI -2.17 to -0.11, p=0.04) were lower in the Pycnogenol^®^ compared to the placebo group. All other outcomes were not different between groups. The proportion of participants reporting fatigue, PEM, dyspnea and brain fog at baseline and follow-up 2 is given in Supplementary Table 2. Mean (standard deviation) unadjusted raw data for the primary and secondary outcomes for both groups are summarized in Supplementary Tables 3-5.

**Table 2.**
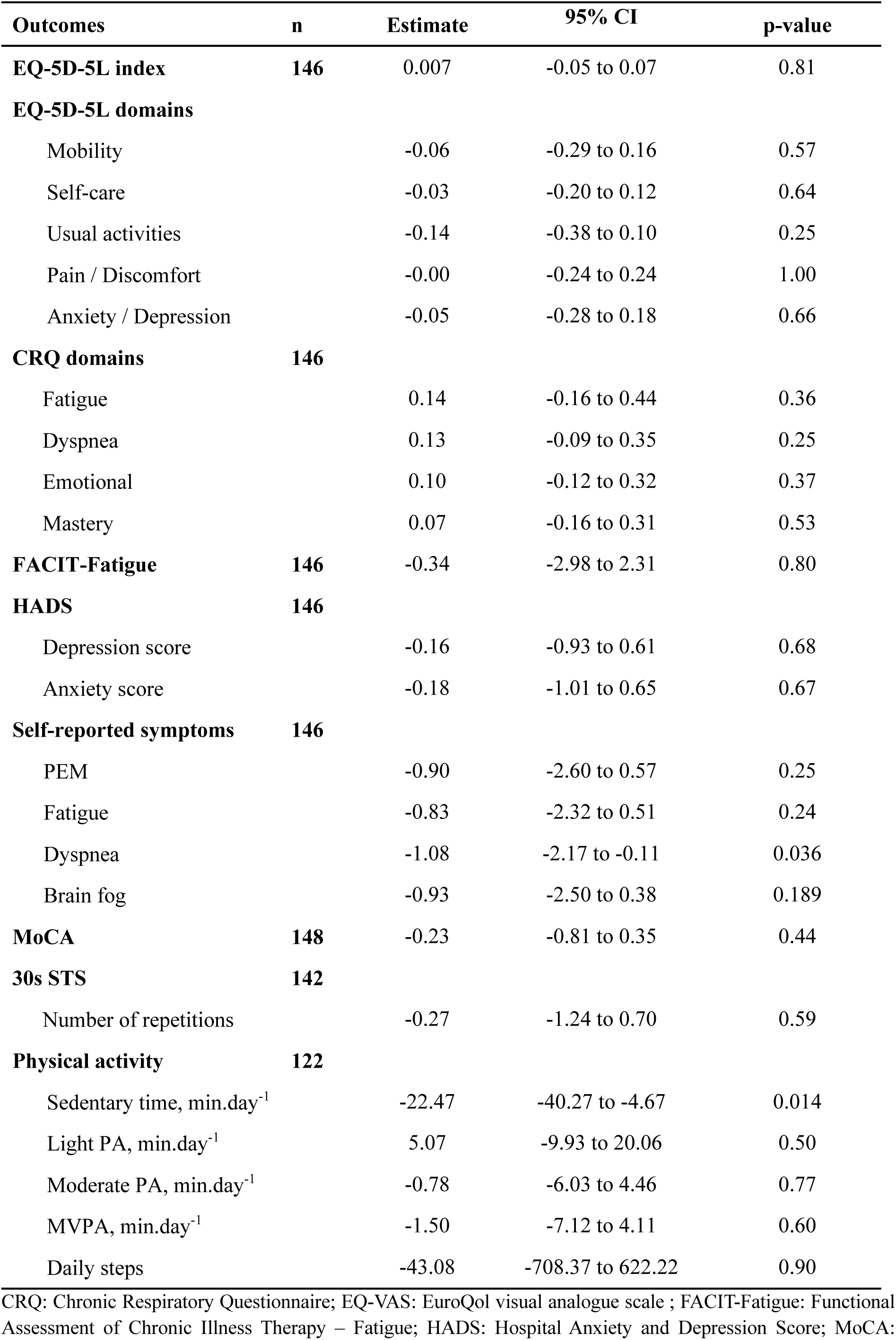
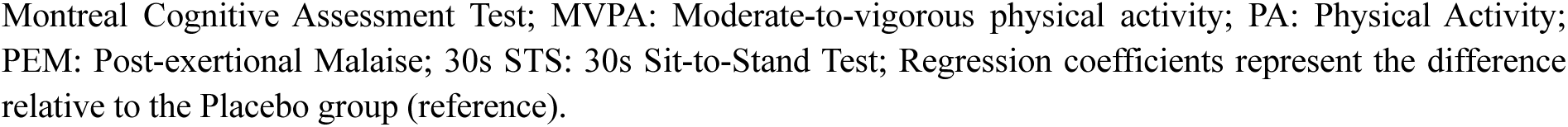
Multivariable linear regression models for secondary outcomes at follow-up 2 (V4), adjusted for mean baseline values and presence of chronic symptomatic disease.

| Outcomes | n | Estimate | 95% CI | p-value |
| --- | --- | --- | --- | --- |
| **EQ-5D-5L index** | **146** | 0.007 | -0.05 to 0.07 | 0.81 |
| **EQ-5D-5L domains** |  |  |  |  |
| Mobility |  | -0.06 | -0.29 to 0.16 | 0.57 |
| Self-care |  | -0.03 | -0.20 to 0.12 | 0.64 |
| Usual activities |  | -0.14 | -0.38 to 0.10 | 0.25 |
| Pain / Discomfort |  | -0.00 | -0.24 to 0.24 | 1.00 |
| Anxiety / Depression |  | -0.05 | -0.28 to 0.18 | 0.66 |
| **CRQ domains** | **146** |  |  |  |
| Fatigue |  | 0.14 | -0.16 to 0.44 | 0.36 |
| Dyspnea |  | 0.13 | -0.09 to 0.35 | 0.25 |
| Emotional |  | 0.10 | -0.12 to 0.32 | 0.37 |
| Mastery |  | 0.07 | -0.16 to 0.31 | 0.53 |
| **FACIT-Fatigue** | **146** | -0.34 | -2.98 to 2.31 | 0.80 |
| **HADS** | **146** |  |  |  |
| Depression score |  | -0.16 | -0.93 to 0.61 | 0.68 |
| Anxiety score |  | -0.18 | -1.01 to 0.65 | 0.67 |
| **Self-reported symptoms** | **146** |  |  |  |
| PEM |  | -0.90 | -2.60 to 0.57 | 0.25 |
| Fatigue |  | -0.83 | -2.32 to 0.51 | 0.24 |
| Dyspnea |  | -1.08 | -2.17 to -0.11 | 0.036 |
| Brain fog |  | -0.93 | -2.50 to 0.38 | 0.189 |
| **MoCA** | **148** | -0.23 | -0.81 to 0.35 | 0.44 |
| **30s STS** | **142** |  |  |  |
| Number of repetitions |  | -0.27 | -1.24 to 0.70 | 0.59 |
| **Physical activity** | **122** |  |  |  |
| Sedentary time, min.day<sup>-1</sup> |  | -22.47 | -40.27 to -4.67 | 0.014 |
| Light PA, min.day<sup>-1</sup> |  | 5.07 | -9.93 to 20.06 | 0.50 |
| Moderate PA, min.day<sup>-1</sup> |  | -0.78 | -6.03 to 4.46 | 0.77 |
| MVPA, min.day<sup>-1</sup> |  | -1.50 | -7.12 to 4.11 | 0.60 |
| Daily steps |  | -43.08 | -708.37 to 622.22 | 0.90 |

CRQ: Chronic Respiratory Questionnaire; EQ-VAS: EuroQol visual analogue scale ; FACIT-Fatigue: Functional Assessment of Chronic Illness Therapy – Fatigue; HADS: Hospital Anxiety and Depression Score; MoCA:
Montreal Cognitive Assessment Test; MVPA: Moderate-to-vigorous physical activity; PA: Physical Activity; PEM: Post-exertional Malaise; 30s STS: 30s Sit-to-Stand Test; Regression coefficients represent the difference relative to the Placebo group (reference).

Figure 3 illustrates the progression of severe symptoms over 12 weeks with baseline and follow-up 2 data from questionnaire responses and symptom diary. This exploratory analysis showed a general decline in symptom severity over time in both groups. The proportion of participants reporting severe symptoms appeared to be higher in the placebo group compared to the Pycnogenol^®^ group, particularly with respect to PEM. None of the symptoms showed significant differences between the groups.

**Figure 3.**
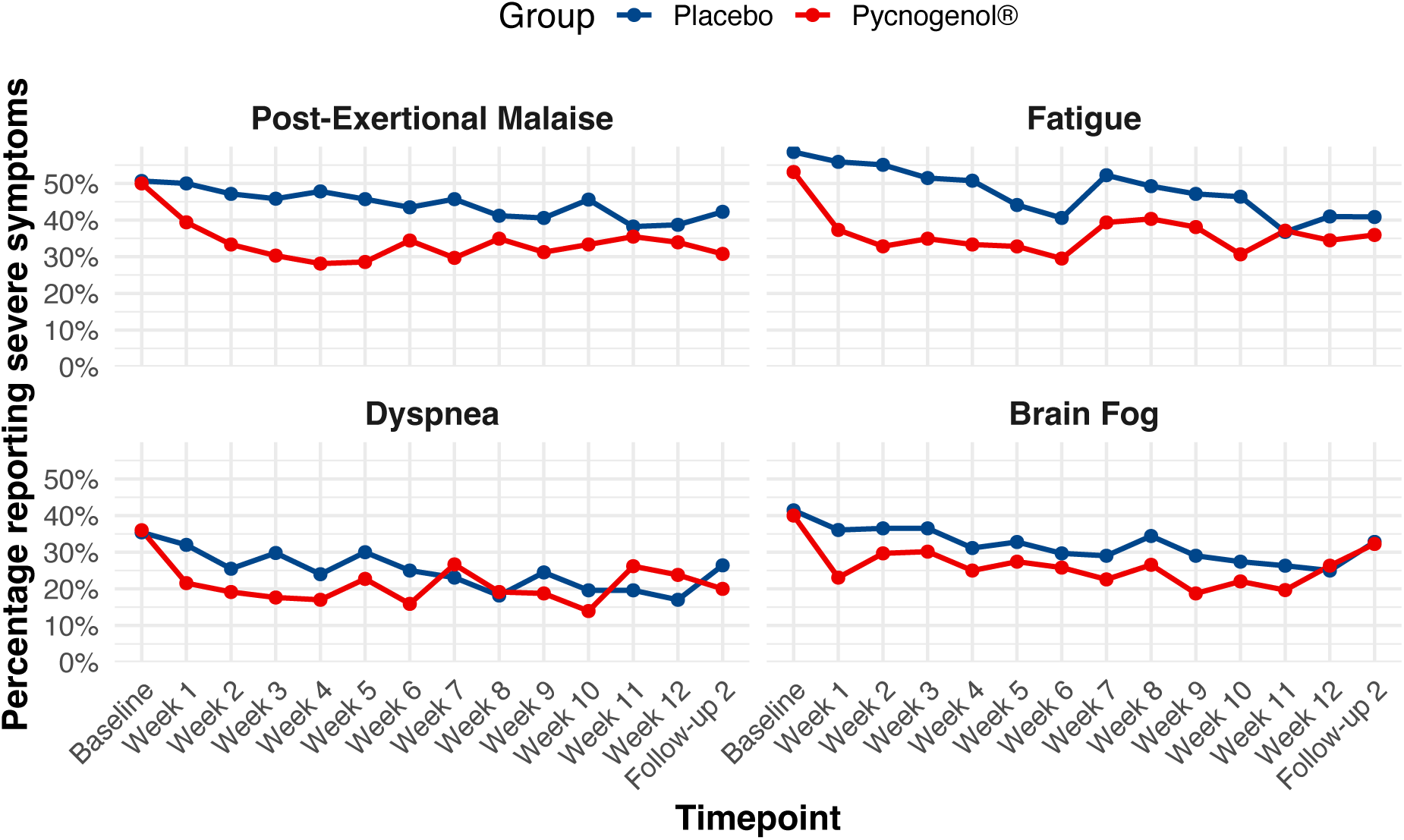
Self-reported severe symptoms over time stratified by randomization group. Dyspnea was assessed using three items: shortness of breath, breathing difficulties at rest, and breathing difficulties during exertion. Brain fog was evaluated based on two items: difficulties with concentration and memory problems. Severity of symptoms was assessed on a 5-point Likert scale ("not bad at all", “mild”, “moderate”, “severe”, and "very severe"). For this analysis, the categories “severe” and “very severe” were combined into one category labelled “severe”. The y-axis represents the percentage of participants with severe symptoms.

Supplementary Figure 3 shows the proportion of participants reporting post-exertional malaise, fatigue, dyspnea, and brain fog at different timepoints for the placebo and Pycnogenol^®^ groups, respectively.

Results for blood biomarkers are summarized in Table 3. Linear regression analysis showed a statistically significant difference in TAC at the measurement timepoint 15min between the Pycnogenol^®^ and placebo group. For all other biomarkers, no significant differences were found.

**Table 3.** Multivariable linear regression models for biomarkers at follow-up 2 (V4), adjusted for mean baseline values and presence of chronic symptomatic disease.

| Outcomes | n | Estimate | 95% CI | p-value |
| --- | --- | --- | --- | --- |
| **Platelet function & endothelium** |  |  |  |  |
| sCD40L, pg/ml | 148 | 0.04 | -0.19 to 0.27 | 0.73 |
| sP-Selectin, pg/ml | 148 | 0.05 | -0.10 to 0.19 | 0.51 |
| sThrombomodulin, pg/ml | 148 | 0.01 | -0.10 to 0.12 | 0.84 |
| VWF, 2.5% plasma | 148 | -0.01 | -0.03 to 0.01 | 0.32 |
| Syndecan-1*, pg/ml | 148 | 0.07 | -0.10 to 0.24 | 0.39 |
| sVCAM-1, pg/ml | 148 | -0.04 | -0.09 to 0.01 | 0.112 |
| **Inflammation & immune response** |  |  |  |  |
| CRP, mg/l | 148 | -0.16 | -0.34 to 0.01 | 0.064 |
| IL-6*, pg/ml | 148 | -0.03 | -0.31 to 0.26 | 0.84 |
| **Antioxidant capacity, mmol/L Trolox-equivalents** |  |  |  |  |
| TAC at 6 min | 148 | 0.01 | -0.00 to 0.02 | 0.132 |
| TAC at 10 min | 148 | 0.01 | -0.00 to 0.02 | 0.077 |
| TAC at 15 min | 148 | 0.01 | 0.00 to 0.03 | 0.048 |
| TAC at 20 min | 148 | 0.01 | -0.00 to 0.02 | 0.059 |
| TAC at 30 min | 148 | 0.01 | -0.00 to 0.02 | 0.105 |

CRP: C-Reactive Protein; IL-6: Interleukin-6; sCD40L: Soluble CD40 Ligand; sP-Selectin: Soluble Platelet Selectin; sThrombomodulin: Soluble Thrombomodulin; sVCAM-1: Soluble Vascular Cell Adhesion Molecule-1; TAC: Total Antioxidative Capacity; VWF: Von-Willebrand-Factor; Regression coefficients represent the difference relative to the Placebo group (reference). \* Considerable proportion of samples below detection limit (Lower Limit of Quantitation (LLOQ) was 8.3 pg/mL for Syndecan-1 and 9.5 pg/mL). A total of 34 samples for Syndecan-1 and 115 samples for IL-6 were below the respective limits of detection.

The per-protocol analysis revealed similar results as the intention-to-treat analysis for the primary and secondary outcomes (Supplementary Tables 6 and 7).

### Safety Outcomes

A total of 31 adverse events (AEs) were recorded in the Pycnogenol^®^ group, of which 11 were classified as probably related to the study product. In the placebo group, 18 AEs were reported, with nine categorized as probably related. Three participants experienced SAEs during the study; all of which were categorized as unrelated to the study product. The most common AEs considered probably related to intake of Pycnogenol^®^ were gastrointestinal and dermatological complaints. Five participants from the Pycnogenol^®^ group and two from the placebo group discontinued the study product due to side effects. Specifically, two participants from the Pycnogenol^®^ group and one from the placebo group withdrew due to gastrointestinal complaints; one from each group due to dermatological complaints; and one participant from the Pycnogenol^®^ group due to other complaints (Supplementary Table 8).

A total of 88 crashes (worsening of symptoms) were reported within three days after the study visit — 46 in the placebo group and 42 in the Pycnogenol^®^ group. Among those, 34 participants in the placebo and 27 participants in the Pycnogenol^®^ group felt that the study visit caused the crashes. One person reported a new symptom after the study visit. Potential reasons for symptom worsening included travel to the study center (reported in 34 cases), physically demanding procedures such as spirometry and the 30-second sit-to-stand test (36 cases), cognitively demanding tasks like the MoCA test or extensive questioning during the visit (39 cases), and individual factors such as prolonged visit duration, blood sampling, or attending the visit in a fasted state (16 cases).

To evaluate the satisfaction of the participants with the study procedures, we invited them to fill-out an online questionnaire after the follow-up 2 visit. Of the 125 participants who filled-out the feedback-questionnaire (83% response rate), 93.5% were satisfied with the organization of the study. Contact with the study team was mostly described as pleasant, friendly, and empathetic. Overall, 96% rated the study team as professional and 97.5% stated that their personal concerns were addressed sufficiently (Supplementary Table 10).

## Discussion

### Principal Findings

Our findings demonstrated that Pycnogenol^®^ was not superior to placebo in improving the health status of persons with PCC. Furthermore, the Pycnogenol^®^ group reported improvements in dyspnea severity, appeared to engage in less sedentary behavior, and had a marginally higher total anti-oxidative capacity.

Perceived health status at baseline was remarkably low in our study population with median EQ-VAS values of about 50 (0-100 scale), highlighting the extent of their perceived health deterioration. Post intervention, we did not observe any differences in perceived health status between the Pycnogenol^®^ and placebo groups. There was considerable intra-individual variability in serial EQ-VAS responses across the seven-day assessment period (Supplementary Figure 2); this approach was chosen deliberately to capture the natural day-to-day fluctuations in health status. A single health status assessment, likely completed on a ’good’ day, may fail to reflect the full spectrum of variability in health status experienced by participants. Therefore, by assessing health status over multiple consecutive days, we aimed to obtain a more comprehensive and accurate picture of participants’ perceived health. However, the magnitude of improvement in median EQ-VAS – 5.4 units in the Pycnogenol^®^ group and 7.9 units in the placebo group – falls within the established minimal important difference of 5-8 units in people with chronic respiratory conditions and may thus be considered clinically relevant (54), regardless of treatment allocation. In the context of PCC, placebo responses and care-related effects may be heightened due to the chronic and often poorly understood nature of symptoms. A recent longitudinal study in people with chronic low back pain demonstrated that higher physician empathy was associated with significantly improved patient-reported pain, function, and quality of life exceeding outcomes of pharmacological and surgical interventions (55). These findings stress the importance of empathy, especially in conditions without clear diagnostic criteria or established therapies. Furthermore, there is growing evidence suggesting that individuals with PCC often experience feelings of being dismissed, stigmatized, or inadequately supported within the healthcare system. A recent meta-synthesis of qualitative studies highlighted that individuals with PCC frequently encounter fragmented care, disbelief from healthcare providers, and difficulty obtaining a diagnosis, especially when objective biomarkers are not available (56). We hypothesize that the observed improvement in health status in our placebo group is partly related to the trial design and staff, also supported by participant feedback. The trial was designed with the support of individuals with lived experience; we offered flexible visits including visits at home, and provided continued care throughout the study which may have positively influenced participant engagement and perceived health, even in the absence of specific pharmacological efficacy. Furthermore, we do not expect that the natural history of the condition to have influenced EQ-VAS during the 12-week study period, as most of our participants had been experiencing PCC for over a year. Improvements in recovery and symptom reduction appear to be less pronounced after 12 months, suggesting that a subset of individuals may transition into a more chronic form of PCC (7).

We did not observe any consistent differences between groups in secondary outcomes including blood biomarkers. Pycnogenol^®^ has been shown to exert anti-inflammatory and antioxidative properties, and to improve vascular endothelial function in different patient groups (23–25). Indeed, we found slightly higher total antioxidant capacity in the Pycnogenol^®^ compared to the placebo group only at time point 15 minutes, although the magnitude of between-group difference is negligible and not clinically relevant. Overall, CRP and IL-6 levels do not indicate a pro-inflammatory state. This may be due to different PCC phenotypes, in which inflammation is not always a key factor, and the relatively long duration of PCC in our cohort. Moreover, many participants had already been taking various medications, nutritional supplements, or natural remedies prior to enrollment (Supplementary Table 1). These included substances with potential anti-inflammatory and antioxidant effects, which may share mechanistic pathways with Pycnogenol^®^. The widespread use of those compounds may have attenuated potential treatment effects and contributed to the lack of between-group differences in patient-reported outcomes and blood biomarkers. Finally, the between group differences in self-reported dyspnea and time spent in sedentary activity favoring the Pycnogenol^®^ group are difficult to attribute to the expected mechanisms of action of the study product.

### Strengths and Limitations

Strengths of this study include its randomized, quadruple-blinded trial design with robust allocation concealment minimizing bias and confounding. Attrition rates were much lower than expected, reflecting the strong engagement of participants throughout the study. We actively maintained contact with participants, ensuring they felt supported and informed, and encouraged them to continue completing questionnaires even if they discontinued the study product. This careful participant follow-up and engagement likely contributed to the low dropout rate and the completeness of our data. Furthermore, we offered home visits to people with more severe symptoms and limited mobility which enabled the inclusion of a broad spectrum of disease severity and likely enhances the generalizability of our findings across the diverse clinical presentation of PCC. To capture patients’ perspectives, we actively involved people with lived experience in the study planning process through a series of three online engagement sessions. Their feedback was used to refine outcome selection, general study procedures, improve acceptability, and ensure that the trial addressed patient-relevant outcomes and concerns (27).

This study has also limitations. At screening, participants were instructed not to change any medications or therapies throughout the study; however, we cannot completely rule out the possibility of background treatments, including new intake of supplements and/or self-initiated interventions. Since the study product was well tolerated and adverse reactions were relatively few in both groups, we expect this to occur in both groups, potentially shifting the observed effects towards the null, i.e., leading to an underestimation of potential treatment effects. Even though stratification was planned based on symptom duration (<6 months vs. ≥ 6 months), only one participant reported persisting symptoms for less than six months prior to enrolment. We therefore did not adjust the models for this variable as initially planned (26). Consequently, our results may not be generalizable to individuals who have only recently started suffering from PCC. Furthermore, the diagnosis of PCC remains challenging, and only few participants had a physician diagnosis. While we requested SARS-CoV-2 infection confirmation, collected detailed information about the onset of persistent symptoms, and consulted treating physicians to exclude other possible diagnoses, we cannot be certain that all our participants had PCC, given the lack of robust diagnostic criteria.

In conclusion, a daily dose of 200mg Pycnogenol^®^ did not improve overall health status compared to placebo in individuals with PCC over 12 weeks. However, both groups showed clinically relevant improvements in their health status highlighting the potential of non-specific therapeutic effects, including care attention, emotional support, and symptom monitoring. The pathomechanisms underlying PCC remain unclear with different clinical phenotypes likely requiring distinct therapeutic approaches. Identifying people for whom Pycnogenol^®^ could serve as a beneficial add-on treatment warrants further investigation.

## Data Availability

Data and data dictionary will be shared upon reasonable request. Data include individual de-identified participant data and de-identified individual responses to self-report assessments. Data request proposals will be overseen by the core study team, and their final decision about data sharing will be binding. Possible data transfers will need to comply with the data transfer agreement guidelines.

## Contributors

JSF, MAP, TR, LK, MR, JK, and AA designed the study and developed the study protocol. JSF is the sponsor of the study and AA is the principal investigator. JK and TR developed the REDCap database. JB performed the sample size calculations and oversaw the statistical analyses. JK is responsible for all statistical analyses. SR and LK were responsible for the laboratory analyses. TR, JK, and MR were responsible for project management and supervision of team members including quality control. JK and TR wrote the first draft of the manuscript. All authors read, revised, and had final responsibility for the decision to submit the manuscript for publication.

## Acknowledgements

We thank Dr Anja Frei (University of Zurich) for supervising the labeling of the study products; Dr Sarah R. Haile (University of Zurich) for preparing the randomization list and implementation into the database; and Dr Babette Winter (University of Zurich) for training and supervision regarding the cognitive assessment test. Furthermore, we thank Dr Tala Ballouz and Dr Dominik Menges (both University of Zurich) for providing health status data from the Zurich SARS-CoV-2 Cohort to enable sample size calculations for this study, and their contributions to study design. We also thank the people with lived experience from the Long COVID Citizen Science Board for their input on study design as well as Altea, Long COVID Switzerland and all physicians who supported us during recruitment.

