## Supplementary Material for "Effects of Pycnogenol^®^ in post-COVID-19 condition (PYCNOVID): A single-center, placebo controlled, quadruple-blind, randomized trial"

Thomas Radtke

Epidemiology, Biostatistics and Prevention Institute

University of Zurich (UZH)

Hirschengraben 84

8001 Zurich, Switzerland

### Methods

Secondary outcomes were assessed prior to the baseline and follow-up 2 visits.

#### *Spirometry*

Spirometry was performed according to the standards of the American Thoracic Society and European Respiratory Society (1) using portable spirometry (PADSY Spiro, Spirosound, Medset Medizintechnik GmbH, Hamburg, Germany). We calculated z-scores using reference equations published by Quanjer et al.(2).

#### *Questionnaires*

Fatigue was assessed using the 13-item Functional Assessment of Chronic Illness Therapy – Fatigue (FACIT-Fatigue) instrument (3). We used a cut-off score of < 34 to report the number (percentage) of participants with fatigue. Norm values from a German population were applied (4). The scale is responsive to change, and triangulated minimal important differences have been established for various patient populations (5). In a small study ($n = 32$) with people attending COVID-19 rehabilitation, the MID for the FACIT-Fatigue scale has been estimated between 2.7 and 3.6 points (5).

Dyspnea (5 items), fatigue (4 items), emotional function (7 items), and mastery (4 items) were assessed with the Chronic Respiratory Questionnaire (CRQ) (6–8). Each item is scored on a 7-point Likert scale (1 = maximum impairment, 7 = no impairment). Domain scores were calculated as the mean of the items in each domain. Higher scores indicate better health status.

Depression and anxiety were assessed using the Hospital Anxiety and Depression Scale (HADS). This scale includes 14 questions with a 4-point Likert-type scale. We used a cut-off value of 7 or higher to report the number of participants with depression and/or anxiety (9).

Quality of life was assessed using the EuroQol EQ-5D-5L with the Dutch value set to calculate EQ-5D-5L index scores (10,11). The questionnaire includes five dimensions on five levels of severity. Higher index scores indicate higher quality of life.

Cognitive function was assessed using the Montreal Cognitive Assessment Test (MoCa). The test is commonly used to evaluate cognitive function in people with coronavirus disease 2019

(COVID-19) and post COVID-19 condition (PCC) (12–15), with a cut-off score < 26 for impairment (13,16). We applied the standard correction for years of education (17).

***Physical activity***

Physical activity was measured with triaxial accelerometer (ActiGraph wGT3X-BT, Pensacola, FL, USA) worn around the hip for eight consecutive days before the baseline and the follow-up 2 visit. The device was programmed to record raw acceleration at a frequency of 30 Hz. Participants were instructed to wear the device during wakening hours, and to remove the monitor during water-based activities.

Raw data files were downloaded using the customer software ActiLife® (Version 6.13.4) and re-integrated into 60s epochs. We checked all raw data files for spurious counts and re-evaluated non-wear time periods. We used the Choi et al. wear time validation algorithm to define time periods continuous zero values (18). No filter was used to analyze the data. Recordings were included in the analysis if they comprised at least four days — including at least one weekend day — with a minimum daily wear time of 10 hours.

We used cut-offs from Troiano et al. (19) to calculate the daily time spent sedentary (0-99 counts.min$^{-1}$), light (100-2019 counts.min$^{-1}$), moderate 2020-5998 counts.min$^{-1}$), and vigorous physical activity (>5999 counts.min$^{-1}$). In addition, the average daily number of steps was calculated.

***Functional exercise capacity***

Functional exercise capacity was assessed using the 30s sit-to-stand test (20) at the screening, baseline and follow-up 2 visits. The first test at the screening visit was done to familiarize study participants with the test procedures. The aim of 30s STS is to perform as many repetitions sit-to-stand repetitions as possible for 30 seconds (20–23). The test was performed on a height adjusted chair (i.e., 90° knee angle), and the same chair height was used for follow-up measurements. Outcome assessors followed standard instructions, and did not verbally encourage study participants during the test. We only counted the sit-to-stand repetitions that were performed correctly (i.e., with fully straightened legs during the sit-to-stand movement and the buttocks touching the chair during the stand-to-sit movement).

***Blood biomarkers***

Biomarkers for inflammation, endothelial injury, coagulation, platelet function, oxidative stress as well as liver and kidney function were measured at the baseline and follow-up 2 visit. Standard diagnostic analyses (haematogram, CRP, international normalized ratio (INR), D-dimers, activated partial thromboplastin time (aPTT), creatinine, aspartate aminotransferase (ASAT), alanine aminotransferase (ALAT) and gamma glutamyltransferase (γ-GT)) were performed by our partner laboratory ANALYTICA Medizinische Laboratorien AG, Falkenstrasse 14, 8024 Zürich. Total antioxidative capacity (TAC) was measured using a colorimetric kit (catalogue number EEA023, Thermo Fisher Scientific, Bleiswijk, the Netherlands) as specified in the manufacturer's protocol. In short, serum samples were diluted 1:2 in PBS and kept on ice prior to the assay. TAC was assessed based on the reduction of the green radical of 2,2′-Azino-bis(3-ethylbenzothiazoline-6-sulfonic acid ($ABTS^{\bullet+}$) to colourless ABTS by serum antioxidants. The Vitamin E analogue Trolox was used as reference and absorbance measured at 414 nm after 6 min, 10 min, 15 min, 20 min and 30 min was converted into Trolox-equivalents. sCD40L, sP-selectin, sThrombomodulin, VWF, Syndecan-1, sVCAM-1 and IL-6 were analyzed with a Luminex 200 bead-based suspension array system and using Invitrogen ProcartaPlex detection kits (catalogue numbers PPX-02-MX7DR74, PPX-04-MX47XM7, EPX040-10825-901, EPX010-12396-901) (Thermo Fisher Scientific, Vienna, Austria) according to the manufacturer's protocol.

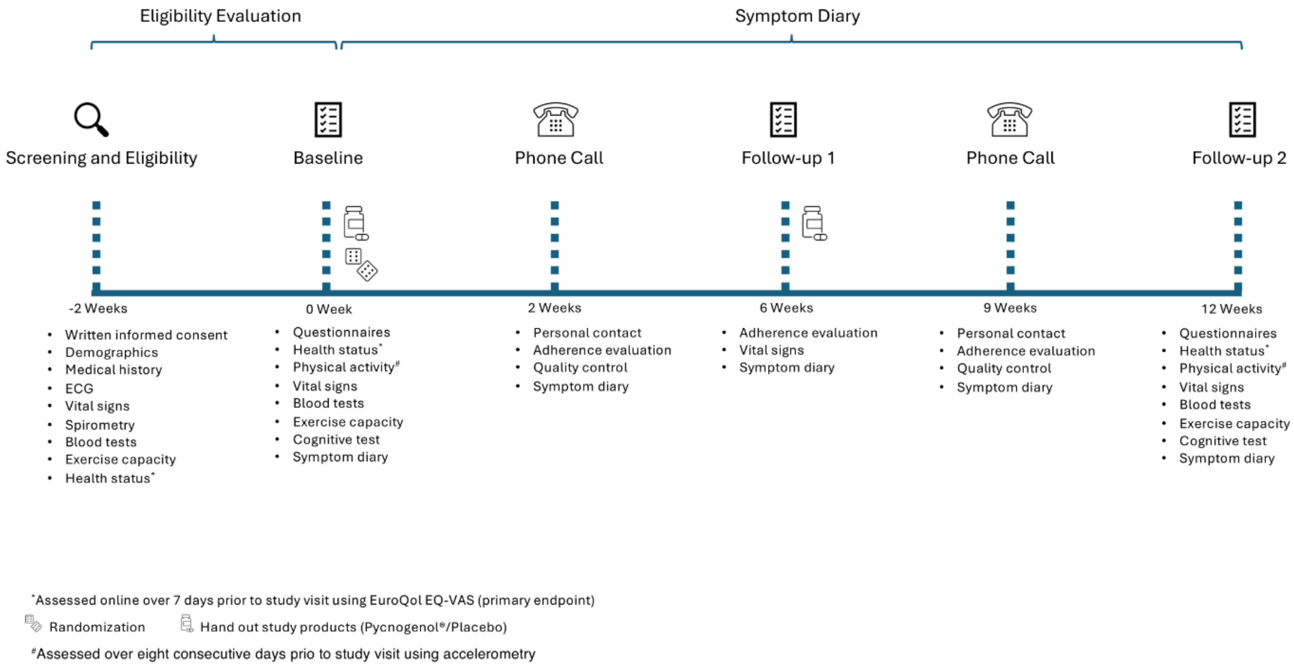


**Supplementary Figure 1.** Timeline of PYCNOVID study assessments and procedures.

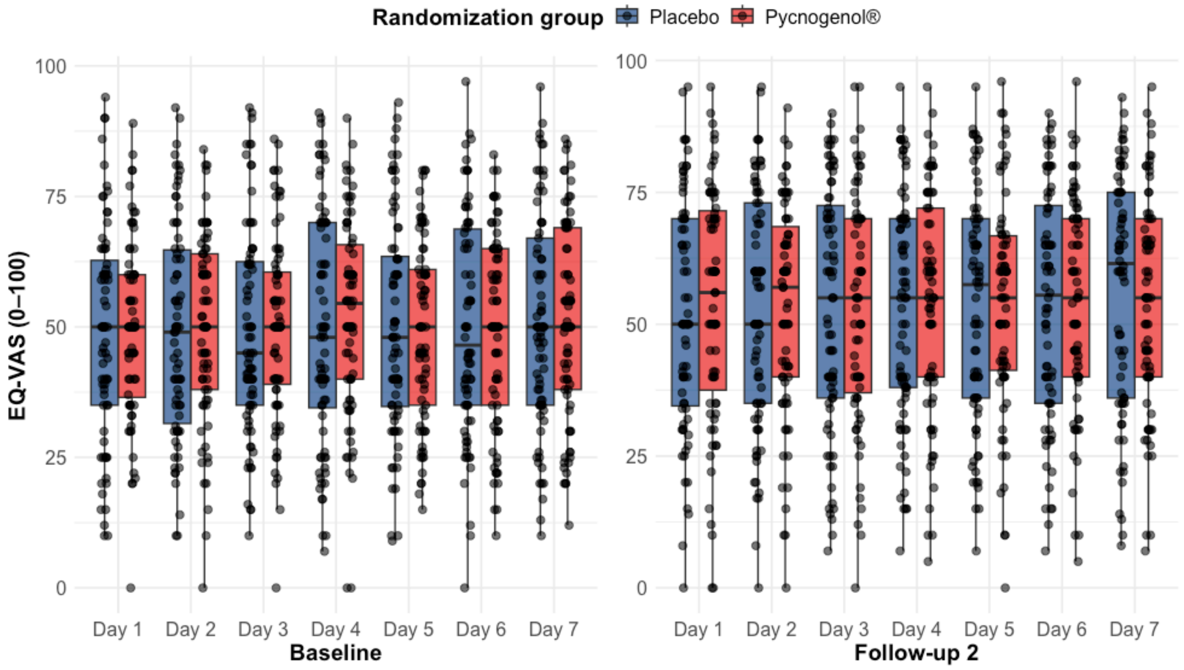


**Supplementary Figure 2.** Raw EQ-VAS values at baseline and follow-up 2 by randomization group. EQ-VAS was assessed daily over seven consecutive days prior to the baseline and follow-up 2 visits, respectively.

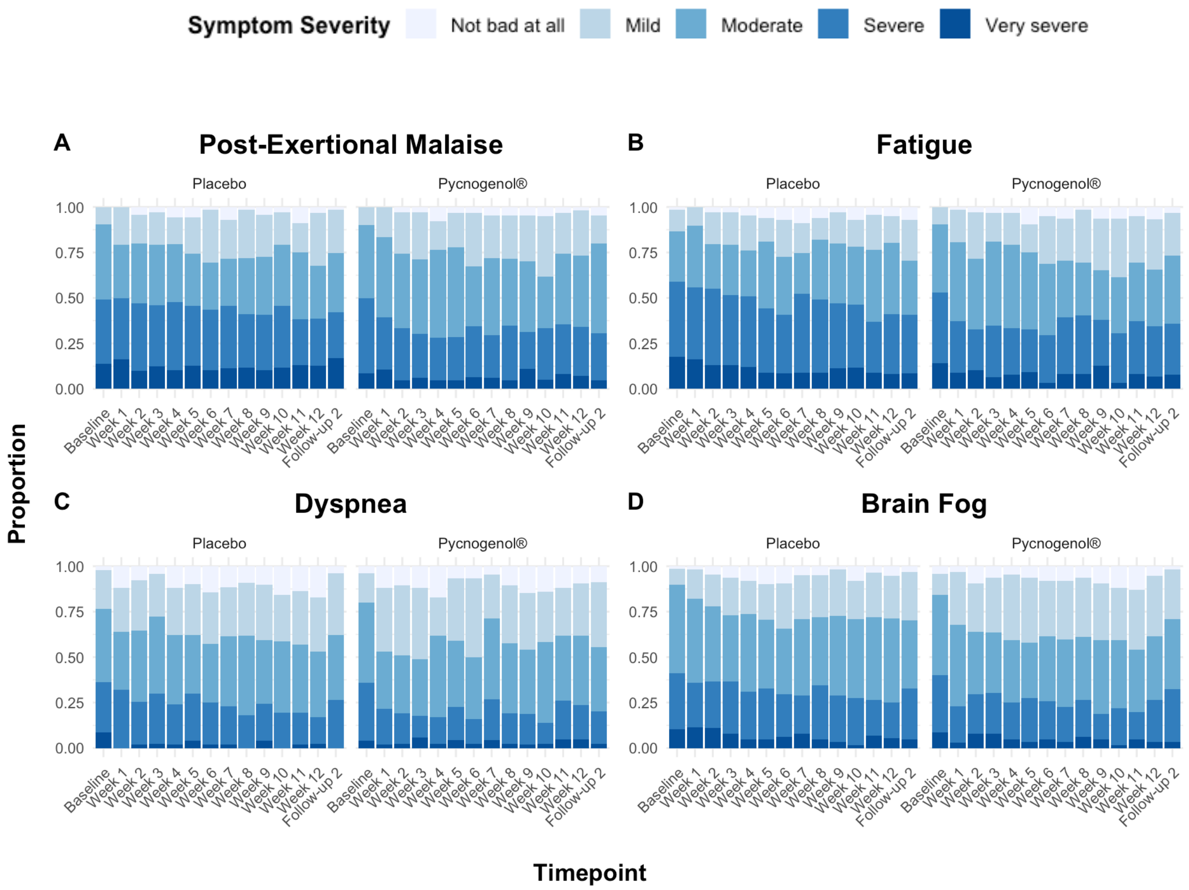


**Supplementary Figure 3.** Symptom severity over time stratified by randomization group. Severity of symptoms was assessed on a 5-point Likert scale ("not bad at all", "mild", "moderate", "severe", and "very severe"). Dyspnea was assessed using three items: shortness of breath, breathing difficulties at rest, and breathing difficulties during exertion. Brain fog was evaluated based on two items: difficulties with concentration and memory problems.

In an exploratory analysis, we examined the effects of time and randomization group on changes in severe symptoms. Participants reporting "severe" or "very severe" symptoms at any timepoint were categorized as "severe"; all others were categorized as "non-severe". We applied a generalized linear mixed-effects model (GLMM) with a binomial family. The model included time and randomization groups as fixed effects, with a random intercept for participants to account for repeated measures. There were no statistically significant differences between the two randomization groups.

**Supplementary Table 1.** Medications and therapies reported by participants for treatment of post-COVID-19 symptoms at baseline.

| **Therapies** | **Placebo n=78** | **Pycnogenol® n=75** |
|---|---|---|
| **No treatment** | 7 (9.0) | 3 (4.0) |
| **Medical therapies incl. medication/supplements** | 48 (61.5) | 41 (54.7) |
| **Prescription medication** | | |
| Phosphodiesterase-4 inhibitors | 0 (0) | 0 (0) |
| Opioid antagonists | 3 (3.8) | 2 (2.7) |
| Muscle relaxants | 3 (3.8) | 4 (5.3) |
| Antihistamines | 14 (17.9) | 16 (21.3) |
| **Medical therapies** | | |
| Apheresis | 3 (3.8) | 2 (2.7) |
| Hyperbaric oxygen therapy | 3 (3.8) | 2 (2.7) |
| **Nutritional supplements** | | |
| Bioflavonoids & Polyphenols | 7 (9.0) | 5 (6.7) |
| Echinacea | 3 (3.8) | 4 (5.3) |
| Ashwagandha | 4 (5.1) | 2 (2.7) |
| Coenzyme Q-10 | 17 (21.8) | 10 (13.3) |
| Padma | 0 (0) | 1 (1.3) |
| Turmeric | 7 (9.0) | 7 (9.3) |
| Black garlic | 1 (1.3) | 1 (1.3) |
| Omega-3 fatty acids | 20 (25.6) | 17 (22.7) |
| Ferritin | 10 (12.8) | 6 (8.0) |
| Zinc | 16 (20.5) | 14 (18.7) |
| Magnesium | 30 (38.5) | 26 (34.7) |
| **Vitamins** | | |
| Vitamin C | 11 (14.1) | 14 (18.7) |
| Vitamin E | 3 (3.8) | 1 (1.3) |
| Vitamin B12 | 17 (21.8) | 12 (16.0) |

| Therapies | Placebo n=78 | Pycnogenol® n=75 |
|---|---|---|
| Multivitamins | 14 (17.9) | 9 (12.0) |
| **Physical activity** | 42 (53.8) | 54 (72.0) |
| **Breathing therapy** | 16 (20.5) | 21 (28.0) |
| **Meditation/Relaxation** | 39 (50.0) | 50 (66.7) |
| **Pacing** | 53 (67.9) | 50 (66.7) |
| **Dietary change** | 16 (20.5) | 29 (38.7) |

Data are reported as numbers (%).

**Supplementary Table 2.** Number (percentage) of the four main symptoms (self-reported) at baseline and follow-up 2 by randomization group.

| | Pycnogenol® n=71 | | Placebo n=75 | |
|---|---|---|---|---|
| **Outcomes** | **Baseline** | **Follow-up 2** | **Baseline** | **Follow-up 2** |
| | N (%) | N (%) | N (%) | N (%) |
| PEM | 67 (94.4) | 64 (90.1) | 71 (94.7) | 71 (94.7) |
| Fatigue | 61 (85.9) | 63 (88.7) | 67 (89.3) | 71 (94.7) |
| Dyspnea | 48 (67.6) | 44 (62.0) | 46 (61.3) | 53 (70.7) |
| Brain fog | 68 (95.8) | 61 (85.9) | 66 (88.0) | 64 (85.3) |

Data are reported as numbers (%). PEM, post-exertional malaise. Severity of symptoms was assessed on a 5-point Likert scale ("not bad at all", “mild”, “moderate”, “severe”, and "very severe"). Self-reported dyspnea was assessed using three items: shortness of breath, breathing difficulties at rest, and breathing difficulties during exertion. Self-reported brain fog was evaluated based on two items: difficulties with concentration and memory problems.

**Supplementary Table 3.** Raw data for the primary outcome, patient-reported outcomes (questionnaires) and functional exercise capacity at baseline and follow-up 2 per randomization group.

| **Outcomes** | **Pycnogenol®** | | | | **Placebo** | | | |
|---|---|---|---|---|---|---|---|---|
| | **Baseline** | | **Follow-up 2** | | **Baseline** | | **Follow-up 2** | |
| | Mean | SD | Mean | SD | Mean | SD | Mean | SD |
| **Primary outcome** | | | | | | | | |
| EQ-VAS, n=150 | 49.87 | 16.08 | 53.92 | 19.60 | 49.41 | 19.58 | 52.95 | 21.41 |
| **Secondary outcomes** | | | | | | | | |
| **EQ-5D-5L index**, n=146 | 0.60 | 0.22 | 0.64 | 0.24 | 0.59 | 0.24 | 0.63 | 0.24 |
| **EQ-5D-5L domains**, n=146 | | | | | | | | |
| Mobility | 1.93 | 0.92 | 1.79 | 0.97 | 1.89 | 1.02 | 1.83 | 0.99 |
| Self-Care | 1.34 | 0.61 | 1.27 | 0.63 | 1.47 | 0.84 | 1.39 | 0.70 |
| Usual Activity | 3.13 | 0.91 | 2.86 | 1.02 | 3.08 | 1.06 | 2.96 | 1.14 |
| Pain/Discomfort | 2.73 | 0.94 | 2.55 | 0.91 | 2.64 | 0.97 | 2.49 | 0.99 |
| Anxiety/Depression | 1.93 | 0.88 | 1.85 | 0.90 | 2.01 | 0.86 | 1.95 | 0.87 |
| **CRQ domains**, n=146 | | | | | | | | |
| Fatigue | 3.31 | 0.88 | 3.73 | 1.14 | 3.62 | 1.10 | 3.82 | 1.25 |
| Dyspnea | 5.70 | 1.23 | 5.86 | 1.21 | 5.63 | 1.24 | 5.66 | 1.18 |
| Emotional | 4.55 | 0.97 | 4.70 | 1.01 | 4.61 | 0.98 | 4.66 | 1.08 |
| Mastery | 5.68 | 0.95 | 5.79 | 1.04 | 5.71 | 0.89 | 5.74 | 0.96 |
| **FACIT-Fatigue**, n=146 | 23.79 | 7.86 | 25.82 | 10.32 | 23.0 | 9.98 | 25.52 | 11.58 |
| **HADS domains**, n=145 | | | | | | | | |
| Depression | 6.59 | 3.52 | 5.80 | 3.77 | 6.61 | 4.05 | 5.97 | 3.92 |
| Anxiety | 5.76 | 3.80 | 5.28 | 4.13 | 5.97 | 3.78 | 5.61 | 3.40 |
| **MoCA**, n=148 | 27.83 | 1.82 | 27.56 | 1.88 | 27.37 | 1.77 | 27.54 | 2.10 |
| **30s STS**, n repetitions, n=142 | 15.04 | 6.17 | 15.80 | 5.97 | 16.49 | 6.95 | 17.29 | 6.96 |

CRQ: Chronic Respiratory Questionnaire; EQ-VAS: EuroQol Visual Analogue Scale; FACIT-Fatigue: Functional Assessment of Chronic Illness Therapy – Fatigue; HADS: Hospital Anxiety and Depression Score; MoCA: Montreal Cognitive Assessment Test; MVPA: moderate-to-vigorous physical activity: PEM: PA: Physical Activity; Post-exertional Malaise; 30s STS: 30s Sit-to-Stand test.

**Supplementary Table 4.** Raw data for physical activity at baseline and follow-up 2 by randomization group.

| Outcomes | Pycnogenol® n=60 | | | | Placebo n=62 | | | |
|---|---|---|---|---|---|---|---|---|
| | **Baseline** | | **Follow-up 2** | | **Baseline** | | **Follow-up 2** | |
| | Mean | SD | Mean | SD | Mean | SD | Mean | SD |
| **Physical activity** | | | | | | | | |
| Wear time, days | 7.75 | 1.2 | 7.18 | 1.0 | 7.53 | 0.9 | 7.44 | 1.1 |
| Wear time, hours.day$^{-1}$ | 860.7 | 71.0 | 836.7 | 70.3 | 831.7 | 63.9 | 839.0 | 89.7 |
| Step count, steps.day$^{-1}$ | 6456.6 | 3060.7 | 6764.1 | 3367.6 | 6083.0 | 2301.4 | 6507.7 | 3367.6 |
| Sedentary time, min.day$^{-1}$ | 581.3 | 89.7 | 555.5 | 76.3 | 569.9 | 79.4 | 572.8 | 90.7 |
| Light PA, min.day$^{-1}$ | 249.8 | 68.7 | 249.4 | 65.1 | 233.1 | 62.2 | 233.8 | 66.0 |
| Moderate PA, min.day$^{-1}$ | 28.8 | 23.8 | 30.8 | 26.5 | 27.0 | 16.7 | 30.8 | 20.1 |
| MVPA, min.day$^{-1}$ | 29.6 | 25.0 | 31.8 | 28.4 | 28.7 | 17.1 | 32.4 | 22.0 |
| Vigorous PA, min.day$^{-1}$ | 0.79 | 2.0 | 0.93 | 2.3 | 0.76 | 2.0 | 1.59 | 4.4 |

MVPA, moderate-to-vigorous physical activity; PA, physical activity, SD, standard deviation.

**Supplementary Table 5.** Raw data for blood biomarkers at baseline and follow-up 2 by randomization group.

| | Pycnogenol® | | | | | Placebo | | | | |
|---|---|---|---|---|---|---|---|---|---|---|
| | **n** | **Baseline** | | **Follow-up 2** | | **n** | **Baseline** | | **Follow-up 2** | |
| | | Mean | SD | Mean | SD | | Mean | SD | Mean | SD |
| **Platelet function & endothelium** | | | | | | | | | | |
| sCD40L, pg/ml | 72 | 66.4 | 77.6 | 71.5 | 74.4 | 76 | 64.2 | 67.8 | 69.7 | 82.2 |
| sP-Selectin, pg/ml | 72 | 180,920 | 145,658 | 182,758 | 152,218 | 76 | 181,963 | 204,656 | 189,756 | 228,006 |
| sThrombomodulin, pg/ml | 72 | 497 | 279 | 509 | 306 | 76 | 618 | 493 | 601 | 441 |
| VWF, 2.5% plasma | 72 | 0.30 | 0.21 | 0.29 | 0.20 | 76 | 0.29 | 0.25 | 0.28 | 0.23 |
| Syndecan-1, pg/ml | 72 | 35.8 | 21.8 | 36.3 | 23.1 | 76 | 44.2 | 49.9 | 42.1 | 41.6 |
| sVCAM-1, pg/ml | 72 | 255,340 | 70,607 | 256,173 | 80,959 | 76 | 252,583 | 64,158 | 263,662 | 72,103 |
| **Inflammation & immune response** | | | | | | | | | | |
| CRP, mg/l | 72 | 2.49 | 4.2 | 1.79 | 3.2 | 76 | 1.16 | 1.4 | 1.86 | 3.5 |
| IL-6, pg/ml | 72* | 7.48 | 7.45 | 7.68 | 5.92 | 76* | 30.1 | 54.0 | 28.9 | 53.1 |
| **Antioxidant capacity,** mmol/L Trolox-equivalents | | | | | | | | | | |
| TAC at 6 min | 72 | 0.43 | 0.04 | 0.43 | 0.04 | 76 | 0.43 | 0.04 | 0.43 | 0.04 |
| TAC at 10 min | 72 | 0.54 | 0.05 | 0.54 | 0.05 | 76 | 0.55 | 0.05 | 0.54 | 0.05 |
| TAC at 15 min | 72 | 0.66 | 0.05 | 0.66 | 0.05 | 76 | 0.66 | 0.05 | 0.65 | 0.05 |
| TAC at 20 min | 72 | 0.75 | 0.05 | 0.75 | 0.05 | 76 | 0.75 | 0.05 | 0.74 | 0.05 |
| TAC at 30 min | 72 | 0.87 | 0.05 | 0.88 | 0.05 | 76 | 0.88 | 0.05 | 0.87 | 0.05 |

CRP: C-Reactive Protein; IL-6: Interleukin-6s; sCD40L: Soluble CD40 Ligand; sP-Selectin: Soluble Platelet Selectin; sThrombomodulin: Soluble Thrombomodulin; sVCAM-1: Soluble Vascular Cell Adhesion Molecule-1; VWF: Von-Willebrand-Factor; TAC: Total Antioxidative Capacity. * Considerable proportion of samples below detection limit (Lower Limit of Quantitation (LLOQ) was 8.3 pg/mL for Syndecan-1 and 9.5 pg/mL). A total of 34 samples for Syndecan-1 and 115 samples for IL-6 were below the respective limits of detection.

**Supplementary Table 6.** Multivariable linear regression models for primary and secondary outcomes at follow-up 2 among participants adhering to the study protocol (i.e., herein defined as >70% intake of investigational products), adjusted for mean baseline values and presence of chronic symptomatic disease.

| Outcomes | n | Estimate | 95% CI | p-value |
|---|---|---|---|---|
| **Primary outcomes** | | | | |
| EQ-VAS | 135 | 0.54 | 3.51 to 4.60 | 0.79 |
| **Secondary outcomes** | | | | |
| EQ-5D-5L index | 132 | 0.02 | -0.04 to 0.08 | 0.44 |
| EQ-5D-5L domains | | | | |
| Mobility | | -0.11 | -0.34 to 0.12 | 0.33 |
| Self-care | | -0.03 | -0.19 to 0.13 | 0.72 |
| Usual activities | | -0.21 | -0.46 to 0.03 | 0.09 |
| Pain/Discomfort | | 0.02 | -0.23 to 0.28 | 0.86 |
| Anxiety/Depression | | -0.08 | -0.33 to 0.17 | 0.52 |
| CRQ domains | 132 | | | |
| Fatigue | | 0.12 | -0.20 to 0.44 | 0.46 |
| Dyspnea | | 0.16 | -0.07 to 0.39 | 0.163 |
| Emotional | | 0.10 | -0.14 to 0.33 | 0.42 |
| Mastery | | 0.11 | -0.14 to 0.36 | 0.38 |
| FACIT-Fatigue | 132 | -0.01 | -2.78 to 2.75 | 1.00 |
| HADS domains | 132 | | | |
| Depression score | | -0.28 | -1.06 to 0.50 | 0.47 |
| Anxiety score | | -0.38 | -1.20 to 0.43 | 0.35 |
| Self-reported symptoms | 132 | | | |
| PEM | | -1.10 | -3.06 to 0.47 | 0.20 |
| Fatigue | | -1.01 | -2.62 to 0.40 | 0.177 |
| Dyspnea | | -1.06 | -2.24 to -0.03 | 0.055 |
| Brain fog | | -0.81 | -2.40 to -054 | 0.26 |
| MoCA | 134 | -0.13 | -0.75 to 0.49 | 0.68 |
| 30s STS, n repetitions | 129 | -0.43 | -1.38 to 0.52 | 0.37 |
| Physical activity | 110 | | | |
| Step count, steps.day$^{-1}$ | | 12.27 | -707.30 to 731.84 | 0.97 |
| Sedentary time, min.day$^{-1}$ | | -21.50 | -40.24 to -2.76 | 0.025 |
| Light PA, min.day$^{-1}$ | | 4.12 | -11.07 to 19.31 | 0.59 |

| | | | |
|---|---|---|---|
| Moderate PA, $min.day^{-1}$ | 0.04 | -5.68 to 5.59 | 0.99 |
| MVPA, $min.day^{-1}$ | -0.79 | -6.88 to 5.30 | 0.80 |

CRQ: Chronic Respiratory Questionnaire; EQ-VAS: EuroQol Visual Analogue Scale; FACIT-Fatigue: Functional Assessment of Chronic Illness Therapy – Fatigue; HADS: Hospital Anxiety and Depression Score; MoCA: Montreal Cognitive Assessment Test; MVPA: moderate-to-vigorous physical activity: PEM: PA: Physical Activity; Post-exertional Malaise; 30s STS: 30s Sit-to-Stand test.

**Supplementary Table 7.** Multivariable linear regression models for blood biomarkers at follow-up 2 among participants adhering to the study protocol (i.e., herein defined as >70% intake of investigational products), adjusted for mean baseline values and presence of chronic symptomatic disease.

| Outcomes | n | Estimate | 95% CI | p-value |
|---|---|---|---|---|
| **Platelet function & endothelium** | | | | |
| sCD40L | 134 | -0.01 | -0.25 to 0.23 | 0.96 |
| sP-Selectin | 134 | 0.08 | -0.06 to 0.23 | 0.26 |
| sThrombomodulin | 134 | -0.00 | -0.12 to 0.12 | 0.96 |
| VWF | 134 | -0.01 | -0.03 to 0.01 | 0.32 |
| Syndecan-1* | 134 | 0.12 | -0.07 to 0.29 | 0.24 |
| sVCAM-1 | 134 | -0.03 | -0.09 to 0.02 | 0.21 |
| **Inflammation & immune response** | | | | |
| CRP | 134 | -0.16 | -0.34 to 0.03 | 0.098 |
| IL-6* | 134 | -0.08 | -0.38 to 0.23 | 0.62 |
| **Antioxidant capacity** | | | | |
| TAC at 6 min | 134 | 0.01 | -0.00 to 0.02 | 0.061 |
| TAC at 10 min | 134 | 0.01 | 0.00 to 0.03 | 0.044 |
| TAC at 15 min | 134 | 0.01 | 0.00 to 0.03 | 0.029 |
| TAC at 20 min | 134 | 0.01 | 0.00 to 0.03 | 0.044 |
| TAC at 30 min | 134 | 0.01 | -0.00 to 0.02 | 0.091 |

CRP: C-Reactive Protein; IL-6: Interleukin-6; sCD40L: Soluble CD40 Ligand; sP-Selectin: Soluble Platelet Selectin; sThrombomodulin: Soluble Trombomodulin; sVCAM-1: Soluble Vascular Cell Adhesion Molecule-1; TAC: Total Antioxidative Capacity; VWF: Von-Willebrand-Factor; Regression coefficients represent the difference relative to the Placebo group (reference). * Considerable proportion of samples below detection limit (Lower Limit of Quantitation (LLOQ) was 8.3 pg/mL for Syndecan-1 and 9.5 pg/mL). A total of 32 samples for Syndecan-1 and 105 samples for IL-6 were below the respective limits of detection.

**Supplementary Table 8.** Adverse events stratified by randomization group and possible relationship to the investigational products.

| Adverse Events Category | Pycnogenol® Related# (n=75) | Placebo Related# (n=78) | Pycnogenol® Not related (n=75) | Placebo Not related (n=78) | SAE Not related |
|---|---|---|---|---|---|
| Gastrointestinal complaints | 7 | 5 | 0 | 0 | 0 |
| Dermatological complaints | 2 | 1 | 0 | 0 | 0 |
| Respiratory tract infections | 0 | 0 | 6 | 2 | 0 |
| Exercise-related accident | 0 | 0 | 1 | 0 | 1 |
| Worsening of pre-existing condition | 0 | 2 | 4 | 0 | 2 |
| Other complaints | 2 | 1 | 9 | 7 | 0 |
| **Total*** | **11** | **9** | **20** | **9** | **3** |

SAE: Serious Adverse Event. # Adverse events that are probably related to either Pycnogenol® or Placebo.

* The total number of events in each group does not correspond to the number of participants who reported an event, as several participants reported more than one event.

**Supplementary Table 9.** Ingredients of study products verum (Pycnogenol®) and Placebo.

| Ingredient | Verum (mg) | Placebo (mg) |
|---|---|---|
| Pycnogenol® (ACTIVE) | 51.5 | 0 |
| Glucidex 12D | 182.5 | 234 |
| Magnesiumstearat | 5 | 5 |
| Talc | 5 | 5 |
| Aerosil 200 VW Pharma | 6 | 6 |
| Total content | 250 | 250 |
| Gelatine caps size 2 (blue color) | 62 | 62 |
| Total weight of the capsule | 312 | 312 |

**Supplementary Table 10.** Feedback of study participants following completion of the study. Responses were categorized by the study team. For Figure 4B, multiple answers were given. Responses were categorized according to the most emphasized response.

| Questions | Responses, n (%) |
|---|---|
| **How did you feel about the study visits?** | |
| Reasonable | 63 (50.8) |
| Pleasant | 29 (17.7) |
| Well reasonable | 22 (17.7) |
| Visit too long | 6 (4.8) |
| Exhausting | 3 (2.4) |
| Repetitive | 1 (0.8) |
| **Did the study visit have an influence on your state of health?** | |
| None | 68 (57.6) |
| Slightly negative | 21 (17.8) |
| Negative | 16 (13.6) |
| Crash | 13 (11.0) |
| **Did you perceive the study team as professional?** | |
| Yes | 116 (95.9) |
| Mostly | 4 (3.3) |
| No | 1 (0.8) |
| **Were your personal concerns as study participant sufficiently addressed?** | |
| Yes | 118 (97.5) |
| No | 3 (2.5) |
| **How did you find the organization of the study?** | |
| Very good | 64 (52.5) |
| Good | 50 (41.0) |
| Potential for improvement | 3 (2.5) |
| Initial difficulties | 3 (2.5) |
| Misleading | 2 (1.6) |
| **How did you perceive the contact with the study team?** | |
| Pleasant | 76 (63.3) |
| Friendly | 26 (21.7) |
| Empathetic | 13 (10.8) |
| Sympathetic | 4 (3.3) |
| Patient | 1 (0.8) |
